# The effect of SGLT2 inhibitor on clinical outcomes in patients with type 2 diabetes and atrial fibrillation

**DOI:** 10.1101/2023.12.19.23300258

**Authors:** Yongin Cho, Sung-Hee Shin, Min-Ae Park, Young Ju Suh, Sojeong Park, Ji-Hun Jang, Dae-Young Kim, So Hun Kim

## Abstract

**Background:** Sodium glucose cotransporter 2 (SGLT2) inhibitors improve clinical outcomes in several populations including type 2 diabetes, chronic renal insufficiency, and heart failure (HF). However, limited data exist on their effects on atrial fibrillation (AF).

**Methods:** We conducted a retrospective cohort study using the National Health Insurance Service database. A total of 4,771 patients with type 2 diabetes and AF who were newly prescribed SGLT2 inhibitors or DPP4 inhibitors were selected and matched in a 1:2 ratio by propensity score with 37 confounding variables. We assessed the effect of SGLT2 inhibitors on the composite outcome of either HF hospitalization or death compared with DPP4 inhibitors.

**Results:** Over a median follow-up of 31 months, patients on SGLT2 inhibitors were associated with a lower risk of hospitalizations for HF or mortality compared to those on DPP4 inhibitors (HR 0.61; 95% CI 0.44-0.85; P=0.004). SGLT2 inhibitor use was also associated with a lower risk of mortality (HR 0.61; 95% CI 0.39-0.94; P=0.025) and CV mortality (HR 0.43; 95% CI 0.21-0.86; P=0.018), but not of MI (HR 1.22 [95% CI 0.72-2.09]; P=0.461) or stroke (HR 1.00 [95% CI 0.75-1.33]; P=0.980). The incidence of hospitalizations for HF, although statistically insignificant, tended to be lower in the SGLT2 inhibitor group (HR 0.63 [95% CI 0.39-1.02]; P=0.062).

**Conclusion:** In a nationwide cohort of patients with type 2 diabetes and AF, SGLT2 inhibitor was associated with a lower risk of mortality compared to DPP4 inhibitor, which may suggest that SGLT2 inhibitors may be considered as the first-line antidiabetic medication in patients with type 2 diabetes and AF.

## Introduction

Atrial fibrillation (AF), the most common sustained cardiac arrhythmia, is known to be associated with higher risks of stroke, heart failure (HF), and mortality.^1^ A meta-analysis has demonstrated that patients with AF had a 4.62-fold higher risk of developing HF compared to those without AF.^2^ Lack of atrial systole and irregular timing of diastolic can lead to elevated left atrial pressures and decreased stroke volume, which can facilitate the occurrence of HF.^3^ Despite the contemporary therapeutic strategies, the burden of AF has remained constant and the risk of mortality in patients with AF has not improved substantially over the last decade.^4^

Recently, sodium glucose co-transporter 2 (SGLT2) inhibitors have been shown to have clinical benefits in patients with type 2 diabetes at high cardiovascular (CV) risk, chronic renal insufficiency, or HF. This includes a reduction of the risk of hospitalization for HF, CV death, and improvement of renal outcomes.^5^ While the mechanisms responsible for the beneficial effects of SGLT2 inhibitors are still under investigation, several mechanisms have been proposed such as reduced preload and afterload through osmotic diuresis and natriuresis, improving vascular function, improving cardiac energy metabolism, preventing inflammation, inhibiting the cardiac Na+/H+ exchange, and increasing erythytropoietin levels.^6^

AF and diabetes often coexist, and when AF is present in patients with diabetes, it can lead to worse clinical outcomes. In a clinical trial with type 2 diabetic patients with established CV disease, those with AF at baseline had higher rates of adverse HF outcomes than those without AF. In this trial, SLGT2 inhibitors reduced HF-related and renal events irrespective of the presence of AF.^7^ Also, several studies have reported that SGLT2 inhibitors reduced the burden of AF or atrial flutter.^8^

However, data on the direct clinical effects of SGLT2 inhibitors in people with AF are still limited. In this study, we evaluate the clinical outcomes of SGLT2 inhibitors compared with DPP4 inhibitors in patients with type 2 diabetes and AF in a nationwide population-based cohort.

## Methods

We analyzed the health records from the Korean Health Insurance Review and Assessment Service database to estimate the effect of SGLT2 inhibitors on clinical outcomes. In this nationwide retrospective observational study, the database covers > 99% of the South Korean population. This contains all health records, including demographics, diagnoses coded with the International Classification of Diseases [ICD]-10, and drug prescriptions.^9^ We used the data from January 1, 2008, to December 31, 2020. Our study protocol was reviewed and approved by the Institutional Review Board (IRB) of Inha University Hospital (2021-11-022), and informed consent was waived by the IRB.

### Study population

We included the patients with both T2DM (ICD-10 code: E11-14 with at least one medication prescription history) and AF (ICD-10 code: I48) who were newly prescribed SGLT-2 inhibitors or DPP-4 inhibitors since 2014, which is when SGLT2 inhibitors were introduced into the South Korean Market. A new user was defined as a patient who had no history of medication of SGLT2 inhibitor or DPP4 inhibitor for at least 1 year and the first prescription date was designated the index date. We excluded the patients if they were diagnosed with chronic kidney disease stages 4 and 5 (ICD-10 code: N18.4-6), or any malignancy (ICD-10 code: C00-97).

### Clinical outcomes

The primary outcome for our analysis was a composite of the first occurrence of either hospitalization for HF (diagnosed as ICD-10 code I50 during the admission) or death. Secondary outcomes assessed included hospitalization for HF, all-cause mortality, CV mortality, stroke, myocardial infarction, a composite of hospitalization for HF or CV mortality, a composite of major cardiovascular events (MACE), defined as death, MI, or stroke, and hypoglycemia (Detailed definitions are described in Appendix 1.). The follow-up period was defined as the period from the index date until the occurrence of any of the clinical outcomes, discontinuation of the index drug, change to comparison drug or combined use, mortality, the latest follow-up date, or the end of the study period (December 31, 2020), whichever occurred first.

### Statistical analysis

Continuous values are presented as means (standard deviations; SD) and categorical values as numbers (percentages). To minimize differences in the baseline characteristics between the two groups, propensity score matching was performed. A total 37 matching variables included age, sex, duration of type 2 diabetes, index year, income, Charlson comorbidity index, prescribed drugs [1 year before the index date, particularly glucose-lowering medications and CVD medications], and comorbidities [1 year before the index date, particularly CVD diseases and microvascular diseases]. The nearest neighbor matching was used with a caliper (0.1). Propensity score matching was performed with a 1:2 ratio. The covariate balances between the two groups were calculated with standardized differences and absolute values < 0.1 (10%) of standardized differences were considered to be non-significant.

After propensity score matching, we performed survival analyses to estimate the effect of SGLT2 inhibitors on clinical outcomes. The Kaplan–Meier estimates were performed. The difference between the survival curves was assessed by the log-rank test. A marginal Cox proportional hazards regression models for a cluster in a matched pair were performed. We assessed the consistency of the drug effect on the primary outcome in subgroups. In addition, to evaluate whether the benefit of the SGLT2 inhibitor varied with the follow-up period after the time of initiation, analyses were performed according to the time after initiation of the drug (30, 90, 180 days, 1, and 3 years after the index date). Two-sided p values < 0.05 were considered statistically significant. All analyses were performed with SAS (ver. 9.4; SAS Institute, Cary, NC, USA) and R software (ver. 4.0.3; R Development Core Team, Vienna, Austria).

## Result

Among 114,166 patients with type 2 diabetes and AF, a total of 23,467 new medication users were included in the cohort analysis (21,816 new users in the DPP4 inhibitor group and 1,616 new users in the SGLT2 inhibitor group). After propensity score matching, 4,771 patients were finally included (Figure 1). Table 1 showed baseline characteristics before and after propensity score matching. The standardized differences in all variables were<0.1 (10%) after propensity score matching, showing the characteristics were well-balanced between the two groups.

**Figure 1.**
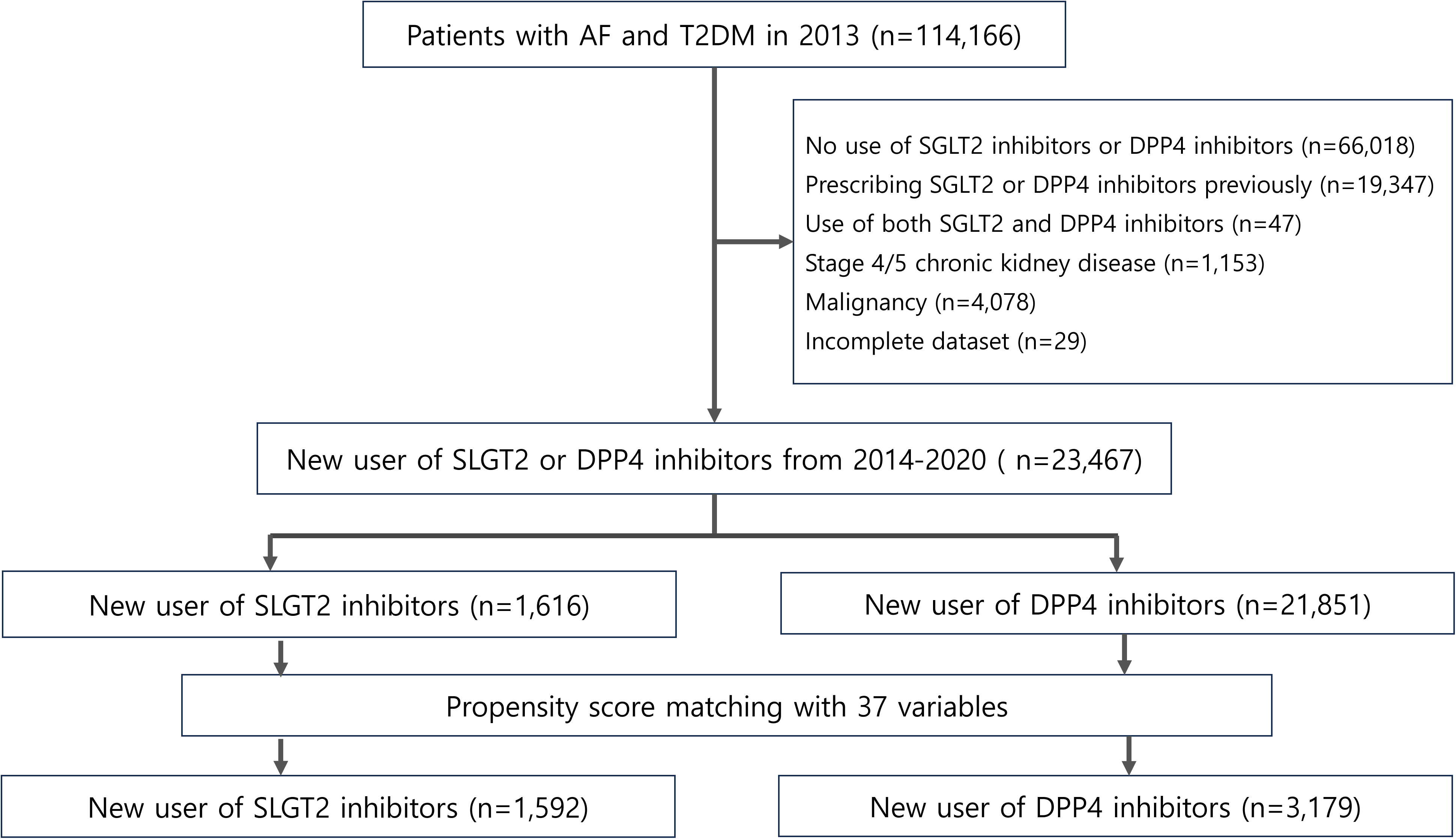
Flow chart of patient selection.

**Table 1.** Baseline characteristics of patients before and after propensity score matching.

|  | Entire population (n=23,467) |  |  |  | Propensity score matched population (n=4,771) |  |  |  |
| --- | --- | --- | --- | --- | --- | --- | --- | --- |
|  | SGLT2i<br>(n=1,616) | DPP4i<br>(n=21,851) | P value | SMD | SGLT2i<br>(n=1,592) | DPP4i<br>(n=3,179) | P value | SMD |
| Mean (SD) age (years) | 67.9 (10.7) | 71.8 (10.1) | <0.001 | -0.380 | 67.9 (10.7) | 68.0 (10.7) | 0.702 | -0.012 |
| Female | 636 (39%) | 9927 (45%) | <0.001 | 0.128 | 623 (39%) | 1261 (40%) | 0.722 | 0.011 |
| Diabetes duration |  |  | <0.001 | 0.745 |  |  | 0.938 | -0.002 |
| ≥6 years | 283 (18%) | 5445 (25%) |  |  | 278 (17%) | 558 (18%) |  |  |
| <6 years | 1333 (82%) | 16406 (75%) |  |  | 1314 (83%) | 2621 (82%) |  |  |
| Index year |  |  | <0.001 | 0.736 |  |  | 0.997 | 0.003 |
| 2014-2015 | 289 (78%) | 10760 (49%) |  |  | 285 (18%) | 567 (18%) |  |  |
| 2016-2017 | 550 (34%) | 6456 (30%) |  |  | 540 (34%) | 1089 (34%) |  |  |
| 2018-2019 | 528 (33%) | 3476 (16%) |  |  | 523 (33%) | 1039 (33%) |  |  |
| 2020 | 249 (15%) | 1159 (5%) |  |  | 244 (15%) | 484 (15%) |  |  |
| Income |  |  | 0.087 | -0.003 |  |  | 0.884 | -0.006 |
| Very low | 357 (22%) | 4866 (22%) |  |  | 357 (22%) | 701 (22%) |  |  |
| Low | 243 (15%) | 2887 (13%) |  |  | 243 (15%) | 475 (15%) |  |  |
| High | 315 (19%) | 4063 (19%) |  |  | 315 (20%) | 660 (21%) |  |  |
| Very high | 678 (42%) | 9717 (44%) |  |  | 677 (43%) | 1343 (42%) |  |  |
| Missing | 23 (1%) | 318 (1%) |  |  |  |  |  |  |
| Charlson comorbidity index |  |  | <0.001 | -0.330 |  |  | 0.922 | 0.012 |
| 0 | 1209 (75%) | 13131 (60%) |  |  | 1187 (75%) | 2387 (75%) |  |  |
| 1 | 131 (8%) | 2335 (11%) |  |  | 131 (8%) | 258 (8%) |  |  |
| ≥2 | 276 (17%) | 6385 (29%) |  |  | 274 (17%) | 534 (17%) |  |  |
| Underlying disease |  |  |  |  |  |  |  |  |
| Hypertension | 1435 (89%) | 18240 (83%) | <0.001 | -0.155 | 1414 (89%) | 2381 (89%) | 0.808 | 0.007 |
| Coronary artery disease | 676 (42%) | 8565 (39%) | 0.037 | 0.056 | 668 (42%) | 1337 (42%) | 0.949 | -0.024 |
| Heart Failure | 655 (41%) | 8124 (37%) | 0.007 | -0.065 | 641 (40%) | 1298 (41%) | 0.707 | 0.012 |
| Stroke | 313 (19%) | 5656 (26%) | <0.001 | 0.154 | 310 (19%) | 614 (19%) | 0.896 | -0.004 |
| Peripheral artery disease | 356 (22%) | 5781 (26%) | <0.001 | 0.108 | 345 (22%) | 751 (24%) | 0.172 | 0.041 |
| Dyslipidemia | 1359 (84%) | 18427 (84%) | 0.803 | 0.009 | 1337 (84%) | 2664 (84%) | 0.872 | -0.005 |
| Chronic kidney disease | 46 (2.9%) | 1415 (6.5%) | <0.001 | 0.113 | 46 (3%) | 112 (4%) | 0.249 | 0.030 |
| Microvascular disease |  |  |  |  |  |  |  |  |
| Diabetic neuropathy | 316 (20%) | 5305 (24%) | <0.001 | 0.113 | 313 (20%) | 639 (20%) | 0.720 | 0.011 |
| Diabetic retinopathy | 326 (20%) | 4888 (22%) | 0.040 | 0.053 | 322 (20%) | 652 (21%) | 0.819 | 0.007 |
| Diabetic nephropathy | 198 (12%) | 3066 (14%) | 0.046 | 0.052 | 196 (12%) | 406 (13%) | 0.652 | 0.014 |
| Glucose-lowering therapies |  |  |  |  |  |  |  |  |
| Insulin | 101 (6%) | 1982 (9%) | <0.001 | 0.108 | 98 (6%) | 199 (6%) | 0.889 | 0.004 |
| Metformin | 1113 (69%) | 15329 (70%) | 0.279 | 0.031 | 1094 (69%) | 2188 (69%) | 0.939 | 0.002 |
| Sulfonylurea | 627 (39%) | 10613 (49%) | <0.001 | 0.197 | 619 (39%) | 1206 (38%) | 0.526 | -0.019 |
| Thiazolidinedione | 261 (16%) | 5094 (23%) | <0.001 | 0.178 | 260 (16%) | 493 (16%) | 0.462 | -0.020 |
| GLP-1 receptor agonist | 124 (8%) | 1428 (7%) | 0.076 | -0.047 | 123 (8%) | 493 (8%) | 0.588 | 0.018 |
| Alpha glucosidase inhibitor | 4 (0.3%) | 12 (0.05%) | 0.004 | -0.050 | 4 (0.3%) | 8 (0.3%) | 0.998 | 0 |
| Cardiovascular therapies |  |  |  |  |  |  |  |  |
| Antithrombotic agents | 1409 (87%) | 18863 (86%) | 0.327 | -0.028 | 1390 (87%) | 2790 (88%) | 0.655 | 0.013 |
| ACE inhibitor or ARB | 1154 (71%) | 14254 (65%) | <0.001 | -0.133 | 1138 (71%) | 2271 (71%) | 0.974 | -0.001 |
| Beta blocker | 919 (57%) | 11294 (52%) | <0.001 | -0.106 | 906 (57%) | 1800 (57%) | 0.850 | 0.013 |
| Calcium channel blocker | 771 (48%) | 10520 (48%) | 0.736 | 0.011 | 757 (48%) | 1532 (48%) | 0.676 | 0.013 |
| Statins | 1178 (73%) | 13921 (64%) | <0.001 | -0.028 | 1161 (73%) | 2323 (73%) | 0.915 | 0.003 |
| Diuretics | 879 (54%) | 12358 (56%) | 0.091 | 0.038 | 871 (55%) | 1689 (53%) | 0.302 | 0.005 |
| Antiarrhythmic drug | 578 (36%) | 8627 (39%) | 0.003 | 0.081 | 565 (35%) | 1156 (36%) | 0.512 | 0.020 |

Values are numbers (percentages) unless stated otherwise.
ACE; angiotensin converting enzyme, ARB; angiotensin receptor blocker, DPP4i; dipeptidyl peptidase 4 inhibitors, GLP; glucagon-like peptide, SD; standard deviation, SGLT2i; sodium glucose cotransporter-2 inhibitors, SMD; standardized mean difference

Over the median follow-up of 31 months (interquartile range, 8 to 75 months), 181 patients (3.7%) died, 70 patients (1.5%) as a result of a CV cause, 121 patients (2.5%) were hospitalized for HF, and 283 patients (5.9%) were hospitalized for HF or died. 95 patients (2.0%) experienced MI, 293 patients (6.1%) had stroke, and 496 patients (10.4%) experienced MACE. In both the Kaplan-Meier and Cox proportional hazards models, patients treated with SGLT2 inhibitor showed a lower risk of HF hospitalization or mortality than those treated with DPP4 inhibitor (HR 0.61 [95% CI 0.44-0.85], P=0.004, Table 2, Figure 2). Additionally, the use of SGLT2 inhibitor was associated with lower risk of mortality (HR 0.61; 95% CI 0.39-0.94; P=0.025), including CV mortality (HR 0.43 [95% CI 0.21-0.86], P=0.018), but not of MI (HR 1.22 [95% CI 0.72-2.09]; P=0.461), stroke (HR 1.0 [95% CI 0.75-1.33], P=0.980), MACE (HR 0.98 [95% CI 0.78-1.23], P=0.831) or hypoglycemia (HR 0.83 [95% CI 0.51-1.35], P=0.446) compared to DPP4 inhibitor. Although the incidence of hospitalizations for HF tended to be lower in the SGLT2 inhibitor group according to Kaplan-Meier analysis (Fig 2), it did not reach a statistically significant level in the Cox regression model (HR 0.63 [95% CI 0.39-1.02]; P=0.062, Table 2).

**Figure 2.**
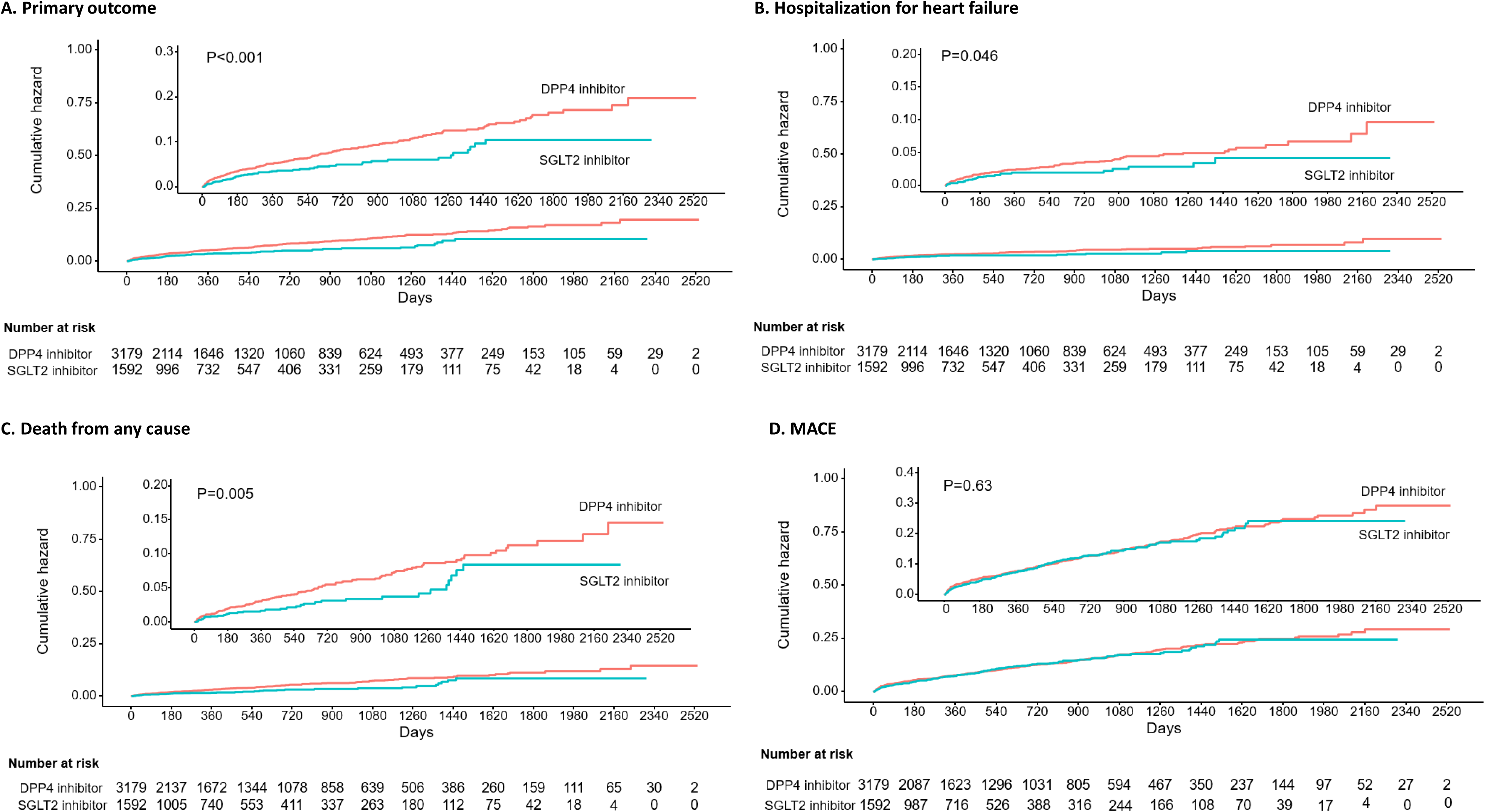
Time to event curves for clinical outcomes. Kaplan–Meier plots of each clinical outcome. A. Primary outcome of hospitalization for heart failure or death from any cause. B. Hospitalization for heart failure. C. Death from any cause. D. A composite of major cardiovascular events. MACE; A composite of major cardiovascular events.

**Table 2.**
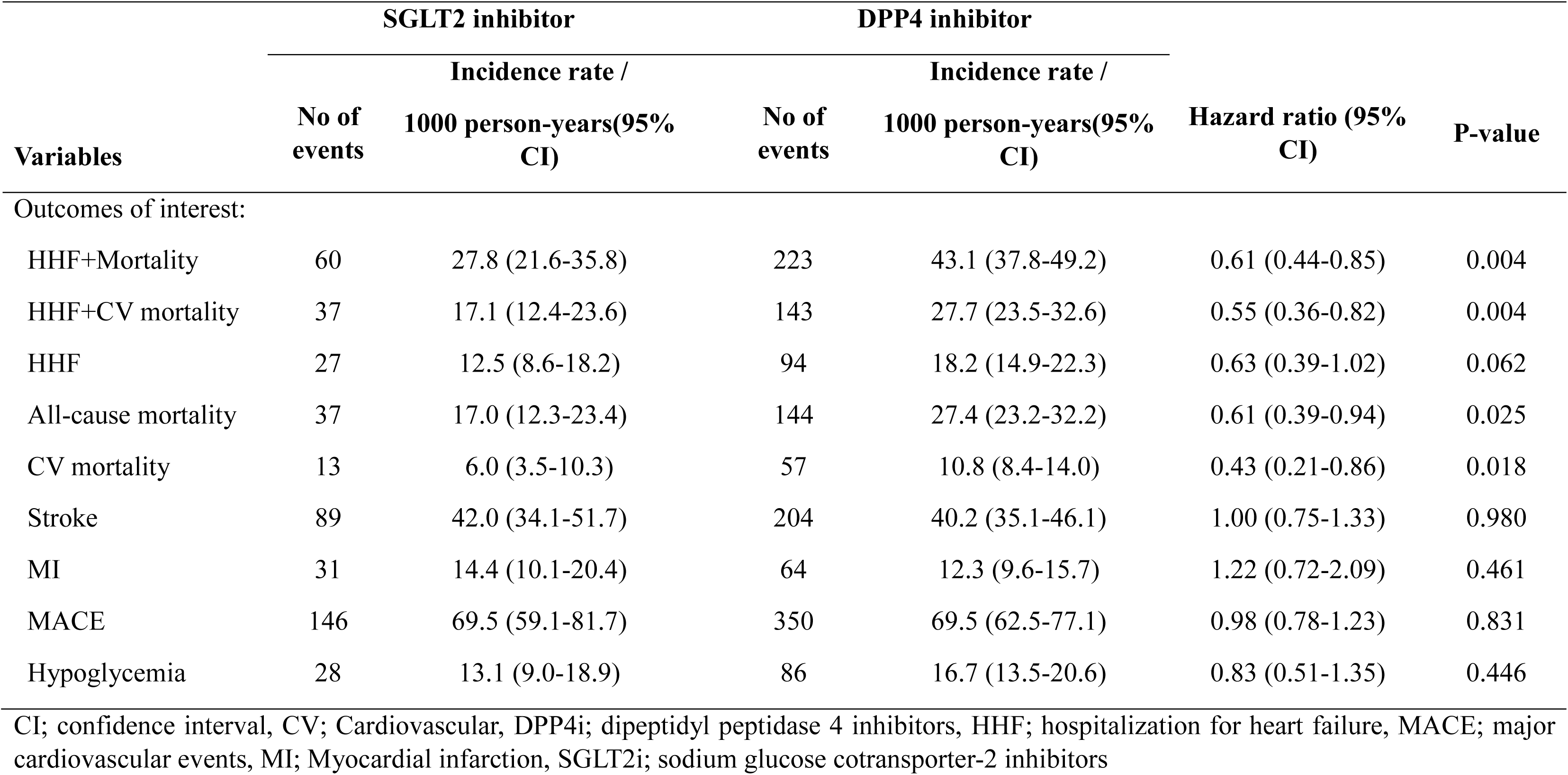
Event rates and hazard ratios for clinical outcomes.

The effect of the SGLT2 inhibitor on the primary outcome was consistent across all subgroups (Figure 3). SGLT2 inhibitors were associated with a lower risk of HF hospitalization or mortality regardless of prior ASCVD or HF history. Patients with SGLT2 inhibitor demonstrated a significantly lower risk of primary outcome from 90 days after initiating the drug to the third year, as compared with those with DPP4 inhibitor (P<0.05, Table 3).

**Figure 3.**
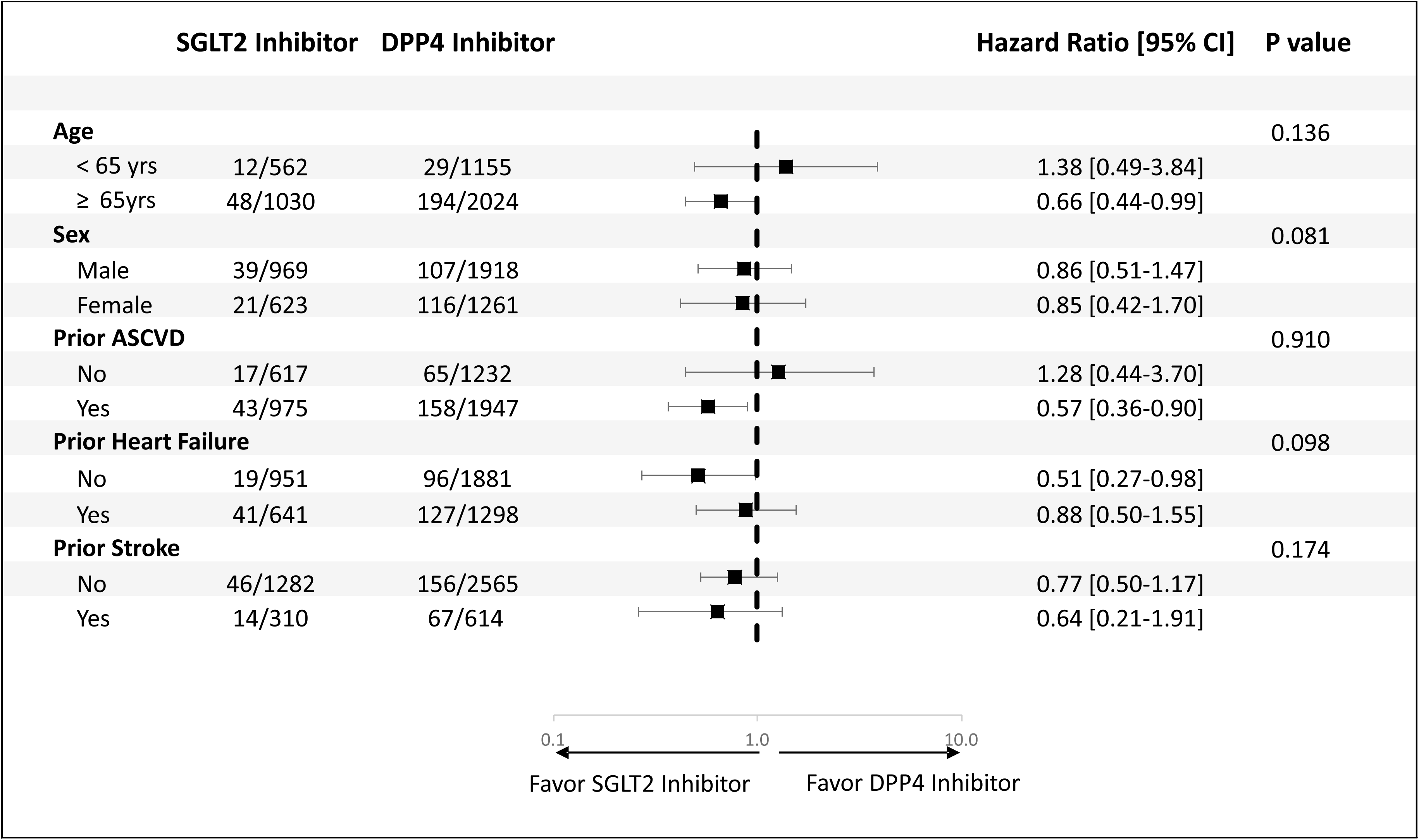
Primary outcomes according to subgroups. Subgroup analysis of forest plot of hazard ratio for SGLT2 inhibitors versus DPP4 inhibitors. Subgroup analysis showed consistent results for a lower risk of the primary outcome of hospitalization for heart failure or death from any cause for SGLT2 inhibitors vs. DPP4 inhibitors (all P interaction > 0.05). HR; hazard ratio, ASCVD; atherosclerotic cardiovascular disease

**Table 3.**
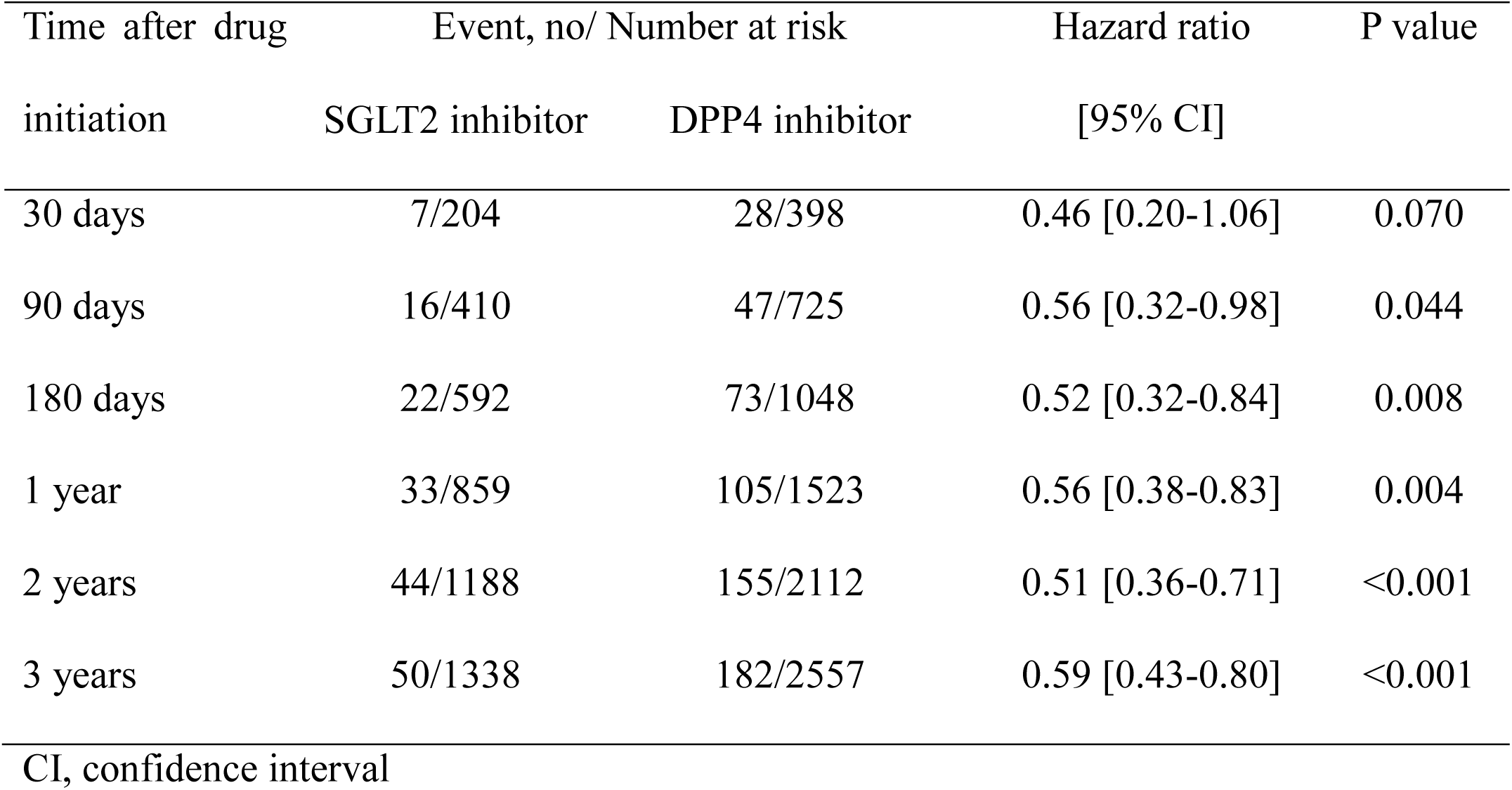
The hazards of hospitalization for heart failure or mortality according to follow-up period.

## Discussion

In this nationwide population-based cohort study, we found that SGLT2 inhibitors were associated with a lower risk of the composite outcome of hospitalization for HF or mortality compared to DPP4 inhibitors in patients with both type 2 diabetes and AF. Use of SGLT2 inhibitors was also associated with a lower risk of mortality including CV mortality, but not with a lower risk of stroke, MI, MACE, or hypoglycemia. The incidence of hospitalizations for HF tended to be lower in the SGLT2 inhibitor group but not statistically significant in the Cox regression model.

In our study, we selected DPP4 inhibitors as an active comparator like other prior studies because it is a relatively new and widely used oral antihyperglycemic agent.^9,10^ Indeed, DPP4 inhibitors are the most commonly used drug in Korea as a second-line therapy after metformin.^11^ In addition, it has shown a neutral effect in most CV outcome trials,^12,13^ and it has a moderate effect in reducing HbA1c with a low risk of hypoglycemia similar to SGLT2 inhibitors.^14^

AF and diabetes are often comorbid conditions, which can be linked through oxidative stress and inflammatory mechanisms. Diabetes can promote the development and maintenance of AF by exacerbating atrial electrical and structural remodeling.^15^ The early stages of diabetes-induced myocardial changes are characterized by increased fibrosis and stiffness, which is reflected by decreased early diastolic filling, increased atrial filling and dilatation, and elevated left ventricular end-diastolic pressure.^16^ Also, in patients with AF, it has been reported that those with diabetes have a worse quality of life and more clinical events including all-cause or CV mortality and hospitalizations, compared to patients without diabetes.^17,18^ Thus, a holistic approach would be important in managing patients with concurrent AF and diabetes.

Clinical trials have shown the beneficial effects of SGLT2 inhibitors in patients with type 2 diabetes at high CV risk or established atherosclerotic CV diseases, HF, or chronic kidney disease. A meta-analysis indicated that SGLT inhibitors resulted in a lower risk in the composite of hospitalization for HF or CV death by 30% in patients with a history of AF, which was similar to the effect estimate for patients without AF.^19^. However, landmark clinical trials included diabetic patients at high risk or specific population, and it may not be generalizable to all the patients with AF in the community. In our study, we demonstrated consistent clinical benefits of SGLT2 inhibitors compared to DPP4 inhibitors in patients with AF and type 2 diabetes using a nationwide cohort with long-term follow-up. And this clinical benefit became apparent from 90 days after initiating the drug. Interestingly, the benefit of the SGLT2 inhibitor was more pronounced for mortality than for HF hospitalization. While all-cause mortality was included as a component of the primary composite endpoint rather than CV mortality in our analysis because there is a potential for misclassification of cause of death due to the characteristics of this nationwide cohort, CV mortality also showed a significantly lower rate in the SGLT2 inhibitor group.

Multiple mechanisms have been proposed for the beneficial effects of SGLT2 inhibitors. SGLT-2 inhibition causes glycosuria, as well as natriuresis, osmotic diuresis, and plasma volume constriction.^20^ This results in beneficial effects on parameters such as glucose concentration, body weight, blood pressure, and albuminuria, which are also associated with reduced risk of CV death and HF.^21^ Recently, other cardioprotective mechanisms have also been discussed. Mechanisms such as prevention of cardiac inflammation and oxidative stress, apoptosis, dysfunction of ionic homeostasis, and mitochondrial dysfunction have been proposed. ^22,23^ These effects could potentially extend to patients with AF. In addition, the reduction of AF burden by SGLT2 inhibitor might be attributable to more favorable clinical outcomes. Several studies have shown that the use of SGLT2 inhibitors was associated with a reduced risk of incident AF. ^24,25^ Osmotic diuretic and natriuresis will cause a reduction in blood pressure and body weight, which subsequently may retard arterial remodeling. Animal study showed SGLT2 inhibitors can ameliorate arterial structural and electrical remodeling through the improvement of mitochondrial function and mitochondrial biogenesis.^26^

Several limitations of our study should be noted. First, the national cohort database we used in our analysis provides drug information only by class for personal information protection. Therefore, it was not possible to assess the effect according to the detailed type of each drug used. Also, detailed patient information or laboratory results were not provided, which did not allow us to conduct analyses related to these aspects. Second, concerns about the risk of HF for some of the DPP4 inhibitors have not been completely resolved.^27,28^ Therefore, the use of DPP4 inhibitors, an active comparator, may have been associated with an increased risk of HF. However, there is no evidence that DPP4 inhibitors are associated with an increased risk of mortality so far. Third, clinical outcomes were ascertained from the national health insurance service database and national statistical database, but not adjudicated from each patient’s medical records or laboratory tests. Despite prior study having shown favorable reliability of primary diagnostic codes of major clinical outcomes in this database, there remains the possibility of misclassification of clinical events. Fourth, we assessed the effects of SGLT inhibitors in diabetic patients with AF. Thus, whether SGLT2 inhibitors improve clinical outcomes in non-diabetic patients with AF is uncertain. Finally, baseline characteristics were different between the SGLT2 inhibitor group and the DPP4 inhibitor group. Although efforts are made to balance clinical characteristics by propensity score matching with 37 variables, residual confounders can still remain.

## Conclusion

In patients with both AF and type 2 diabetes, SGLT2 inhibitors were associated with a lower risk of either hospitalization for HF or mortality compared to DPP4 inhibitors, which may suggest that SGLT2 inhibitors may be considered as the first-line antidiabetic medication in patients with AF and diabetes.

## Declarations

### Ethical Approval

This study complied with the Declaration of Helsinki and was approved by the Institutional Review Board (IRB) of Inha University Hospital (2021-11-022), and informed consent was waived by the IRB.

## Availability of data and materials

The data supporting the findings of this study are accessible from the Korea National Health Insurance Service, but availability is restricted as these data were utilized under license for the current study and are not publicly accessible. However, the data can be obtained from the Korea National Health Insurance Service upon reasonable request and following the appropriate review process.

## Disclosure

Minae Park and Sojeong Park are employees of the Hanmi Pharaceutical Co. They independently performed statistical analysis of the work as coauthors. All other authors have no potential conflict of interest to disclose.

## Author Contributions

Conceptualization: Y Cho, S-H Shin, SH Kim.

Data curation: S-H Shin, M-A Park, S Park.

Formal analysis: M-A Park, S Park.

Investigation: Y Cho, S-H Shin, YJ Suh, J-H Jang, D-Y Kim, SH Kim.

Project administration: S-H Shin, SH Kim.

Software: M-A Park, S Park.

Validation: Y Cho, S-H Shin, SH Kim.

Writing - original draft: Y Cho, S-H Shin

Writing - review & editing: Y Cho, S-H Shin, SH Kim.

---

**Appendix 1.** The definitions and codes used for each outcome.

| **Outcomes** | **Logical description and ICD-10 codes** |
| --- | --- |
| Hospitalization for heart failure | Admission with the following ICD-10 codes: I50<br>Visit ER and diuretics treatment with the following ICD-10 codes: I50 |
| All-cause mortality | All death confirmed by death certificate |
| Cardiovascular mortality | Cardiac death confirmed by death certificate.<br>Death from ICD-10 codes I00-I99 |
| Stroke | Admission with the following ICD-10 codes: I60, I61, I63, I64, G45, or H34 |
| Myocardial infarction | Admission with the following ICD-10 codes: I21, I22 |
| A composite of major cardiovascular events | All death, myocardial infarction or ischemic stroke |
| Hypoglycemia | Admission with the following ICD-10 codes: E10.0, E11.0, E11.63, E12.0, E13.0, E14.0, E16.0-2 |

